# Application of decision-analytic models to inform integrated care interventions for cardiometabolic multimorbidity: A systematic review

**DOI:** 10.1101/2024.10.19.24315798

**Authors:** Elvis O. A. Wambiya, Duncan Gillespie, Robert Akparibo, James O. Oguta, Catherine Akoth, Peter Otieno, Peter J. Dodd

## Abstract

**Introduction:** Integrated care is increasingly being adopted to address the complex needs of patients with cardiometabolic multimorbidity. However, it is unclear how to cost-effectively configure health service pathways for these patients. This study aimed to review and appraise decision analytic models (DAMs) used in economic evaluations of integrated care interventions for patients with cardiometabolic multimorbidity.

**Methods:** We conducted a systematic search for peer-reviewed articles in eight electronic databases, published in English language until December 2023. Any study worldwide that used a decision-analytic model to conduct an economic evaluation of an integrated care model for patients with cardiometabolic multimorbidity was included. We summarised characteristics of the DAMs, integrated care models evaluated, diseases constituting multimorbidity, and critically appraised the quality of reporting of the economic evaluations using Philips (2006) checklist.

**Results:** Out of 16 model-based assessments of the differences between alternative integrated care pathways, most studies (n=13, 81%) were cost utility analyses, focused on care for patients with hypertension and/or diabetes concordant multimorbidity (n=11, 69%). Most studies were conducted in high-income countries (n = 11, 69%). More than half (n = 10, 63%) of the studies used simulated Markov models, while only three studies used individual sampling (microsimulation) models. Few studies were explicit about their data validation approaches against local data, quality of data incorporated in the models, and internal and external consistency.

**Conclusion:** Decision-analytic models investigating integrated care pathways for cardiometabolic multimorbidity should employ microsimulation to describe and incorporate repeated patient interactions with health care and multimorbidity outcomes in the economic evaluations. Consideration of uncertainty in data sources and model structure is also needed to provide robust conclusions. The study also highlighted the need for more economic evaluations using DAMs in low- and middle-income countries to evaluate integrated care models in the context of cardiometabolic multimorbidity.

## What is already known?

- Integrated care is an effective and recommended intervention in improving health outcomes for people with multimorbidity.
- Decision-making on the most cost-effective configuration of integrated care for cardiometabolic multimorbidity is complex because there are so many ways to integrate care to fit specific health services provision and population contexts.
- Model-based explorations of the alternative service specifications of integrated care are useful in tailoring them to the target populations in the most cost-effective way and therefore informing health care decision making.

## What are the new findings?

- This systematic review synthesised evidence on the application of decision-analytic models (DAMs) used in economic evaluations of integrated care interventions in the context of cardiometabolic multimorbidity.
- Most of the DAMs evaluating integrated care are Markov models, performed from a health system perspective, and considering hypertension or diabetes as the main disease conditions.
- There is limited evidence is the application of DAMs evaluating integrated care models for cardiometabolic multimorbidity from low- and middle-income countries (LMICs).

## What do the new findings imply?

- The complexity of integrated care in the context of multiple diseases indicates the need for more studies using individual patient simulation models that can better describe repeated interactions with health care for patients with multimorbidity.
- Future DAMs should be more transparent in their considerations and reporting of data incorporation, assessment of uncertainty, model validation, internal and external consistency.
- More studies using DAMs to evaluate integrated care for cardiometabolic multimorbidity are needed to inform decision making in LMICs.

## Background

Chronic diseases are generally defined as conditions lasting at least a year and requiring continued medical attention or limit activities of daily living or both ^1^. They are a leading cause of morbidity, mortality, and disability globally, making them an important focus for health systems ^2–5^. Recent evidence highlights an increasing global burden of chronic diseases which is attributable to socio-demographic and lifestyle changes, and increased life expectancy due to improved therapies ^6–8^. This increase has contributed to the growing number of people living with multiple chronic conditions making multimorbidity, the simultaneous existence of two or more chronic diseases in an individual, a pertinent public health topic ^9–11^. Multimorbidity is associated with increased disability, morbidity and mortality, reduced quality of life, and polypharmacy leading to adverse drug reactions ^12–16^. In addition, it results in higher health care costs to the patients affected and the health system ^17–19^. Given the complex array of different types of multimorbidity, the contexts of the individuals’ lives and the services that are involved in treating it, understanding how to cost-effectively improve services that treat multimorbid patients is a major challenge for health systems.

Multimorbidity carries a significant burden globally and the distribution and patterns vary across populations, geographical areas, and health care settings ^20–22^. A recent meta-analysis of 68 community-based studies among people aged 45 years and above estimated that at least a third of these populations have two or more chronic diseases with the prevalence being higher in high income-(HICs) than low-and middle-income countries (LMICs) ^23^. Cardiometabolic multimorbidity is considered one of the most common types of multimorbidity ^24–26^. In LMICs however, there is a rising burden of multimorbidity linked to the changing disease landscape characterised by a rise in chronic non-communicable disease burden in the context of persistent chronic communicable diseases ^24,27,28^. This inevitably puts pressure on the health systems of these countries which have been primarily designed to address acute episodic care leading to fragmentation of health services, yet patients with multimorbidity have higher utilisation and complex needs for the comorbidities and their complications ^18,29–33^. Due to the complex nature of multimorbidity, a more comprehensive approach to service delivery that transcends beyond a single-disease focus and is person-centred is needed. However, this would likely require large scale system change to services that would in turn require careful consideration of the relative cost-effectiveness of the alternative options.

Integrated care has been widely adopted to reduce fragmentation and promote comprehensive delivery and promote efficiency of health services ^34^. Traditionally more prominent in HICs, integrated care is increasingly gaining prominence in LMICs to address unique health challenges faced by people with multiple chronic diseases ^35–37^. Different integrated care models have been developed for health care delivery in diverse service contexts to meet the needs of patients with multimorbidity ^38–40^. Existing evidence, mainly from trial settings, demonstrates the effectiveness of integrated care in improving access to and utilisation of care, quality of care, service delivery, clinical outcomes, and cost-saving for people with multimorbidity ^41–45^. The diversity of integrated care models and limited evidence from real-world studies presents a challenge for economic evaluations aimed at decision-making on integrated care and indicates the need for model-based appraisals of alternative options for integrated care tailored to specific service and population contexts.

Decision analytic models (DAMs) provide a systematic approach to evaluate the impact of health interventions on costs and outcomes under alternate scenarios ^46^. They use mathematical relationships to define a series of possible consequences that would occur from a set of alternatives being evaluated, and can be implemented through different model-based approaches ^47–49^. DAMs are particularly suited to addressing the decision-making challenges in integrated care as they enable the flexible specification of the population, disease mechanisms and diverse intervention components, allowing the computation of cost-effectiveness metrics that allow comparisons of different specifications of integrated care in the context of multimorbidity. Although some studies have been published using DAMs to model the impact of integrated care for people with multiple diseases in diverse settings ^50–52^, there has been no attempt at synthesising the modelling approaches taken in a systematic review to understand and appreciate the breadth and quality of evidence. Therefore, the present systematic review aimed to answer the question, how have DAMs been applied to evaluate the health economic impact of integrated care models for patients with cardiometabolic multimorbidity? The aim was to assess the suitability of the DAMs found for assessing the cost-effectiveness of integrated care in the context of cardiometabolic multimorbidity. Based on the review results, we provide recommendations for best practice in decision-analytic modelling for decision making regarding integrated care models for people with cardiometabolic multimorbidity.

## Methods

The systematic review followed methods specified in a registered protocol on PROSPERO (CRD42023407278) ^53^. The findings of this systematic review are reported following the Preferred Reporting Items for Systematic Reviews and Meta-Analysis (PRISMA) guidelines^54,55^.

### Search strategy and literature search

A systematic literature search was conducted in eight electronic peer-reviewed databases including Medline, Web of Science, EMBASE, Cumulative Index to Nursing and Allied Health Literature (CINAHL), APA Psychinfo, Econlit, Scopus, and the Cochrane register of controlled trials between 20/11/2023 and 15/12/2023, in English language, without limits in the time frame. The search strategy captured four key concepts: 1) model-based health economic evaluations 2) integrated care 3) chronic diseases and 4) cardiometabolic diseases. The search strategy was initially piloted in Medline, Embase and Web of Science, and was refined with the help of an information specialist and adapted for each specific database. Full search terms are available in online supplemental appendix section S1.

### Inclusion criteria

Eligible studies included economic evaluations reporting cost-effectiveness or cost-utility (where the outcomes are measured in quality-adjusted life years) outcomes ^56,57^. Descriptive studies, opinion pieces, conference or dissertation abstracts and protocols were excluded. We defined cardiometabolic multimorbidity as the existence of two or more chronic diseases in the same individual, at least one of which was a cardiometabolic disease. Concordant multimorbidity is defined as the co-existence of two or more chronic diseases all of which are cardiometabolic diseases, while discordant cardiometabolic multimorbidity is existence of two or more chronic diseases at least one of which is a cardiometabolic ^58,59^. Integrated care was defined as health service delivery containing two or more components of the chronic care model (CCM), and at least one element of Singer et al.’s, (2011) ^60^ framework for measuring integrated patient care for patients with multiple or complex chronic conditions. Studies not published in the English language, and review papers were excluded. We checked through the reference list of review papers to identify potentially relevant studies that met our inclusion criteria. Definition of terms used for this review are outlined in online supplemental appendix table S1. The inclusion and exclusion criteria summarised using the population, intervention, comparator, outcome (PICO) framework are presented in online supplemental appendix table S2.

### Study selection

The studies identified by the searches were independently screened by three reviewers. Using a predefined selection checklist, the reviewers first screened the titles and abstracts and those that met the eligibility criteria proceeded to full text screening. Reviewers were blinded to each other’s decisions throughout the screening process and any conflicts identified from the screening were resolved through discussion with a fourth reviewer (either RA, DG, or PD). Endnote software was used for the removal of duplicates while Covidence software aided the screening. A detailed explanation of the process is provided in online supplemental appendix section S2.

### Data extraction

Data was extracted electronically by two reviewers using a pre-specified Miscrosoft Excel spreadsheet. The data extraction tool was piloted to ensure that it captured all the required information based on the review objectives. The data that was extracted from the selected studies pertained to study characteristics (study title, authors, year of publication, study setting, study aim, target population), details regarding the decision-analytic model (model type/ approach, integrated care model/intervention evaluated, comparators, model assumptions, model inputs and their sources, multimorbidity conditions modelled, disease parameters included, model limitations), results and conclusions of the study. The findings are reported on a summary table and further described narratively.

### Quality assessment

We used the Philips et al., (2006) checklist to evaluate the quality of the included studies in three domains: structure, data inputs, and consistency ^61^. The assessment was completed by one reviewer (EW) and validated by at least one of the co-authors (JO, DG, RA, or PD). For each item on the checklist, a value of “yes,” “No”, “unclear” or “not applicable” was attributed, which corresponded to numeric values of 1, 0, and 0.5 respectively. We then calculated a mean quality score for each study and for each item across the studies.

### Data synthesis

We undertook a narrative synthesis of the data to summarise and appraise the identified model-based economic evaluations of integrated care for cardiometabolic multimorbidity. We first summarised the population and integrated interventions modelled by the geographical distribution, integrated care model types, and disease conditions modelled. Secondly, we described the decision-analytic approaches used including the type of DAMs, model perspectives, model horizon, and model adaptations which are important aspects in the modelling of integrated care. Finally, appraised the quality of the DAMs used for the economic evaluation of integrated care using the Philips (2006) checklist ^61^. This process enabled the critical synthesis of the considerations in the use of DAMs in economic evaluations of integrated care for cardiometabolic multimorbidity. We have presented results of the economic evaluations which may be useful for readers in online supplemental appendix section S3 and online supplemental appendix table S4.

## Results

The results of the study selection are presented in the PRISMA flow chart (Figure 1). We ultimately included 16 articles.

**Figure 1:**
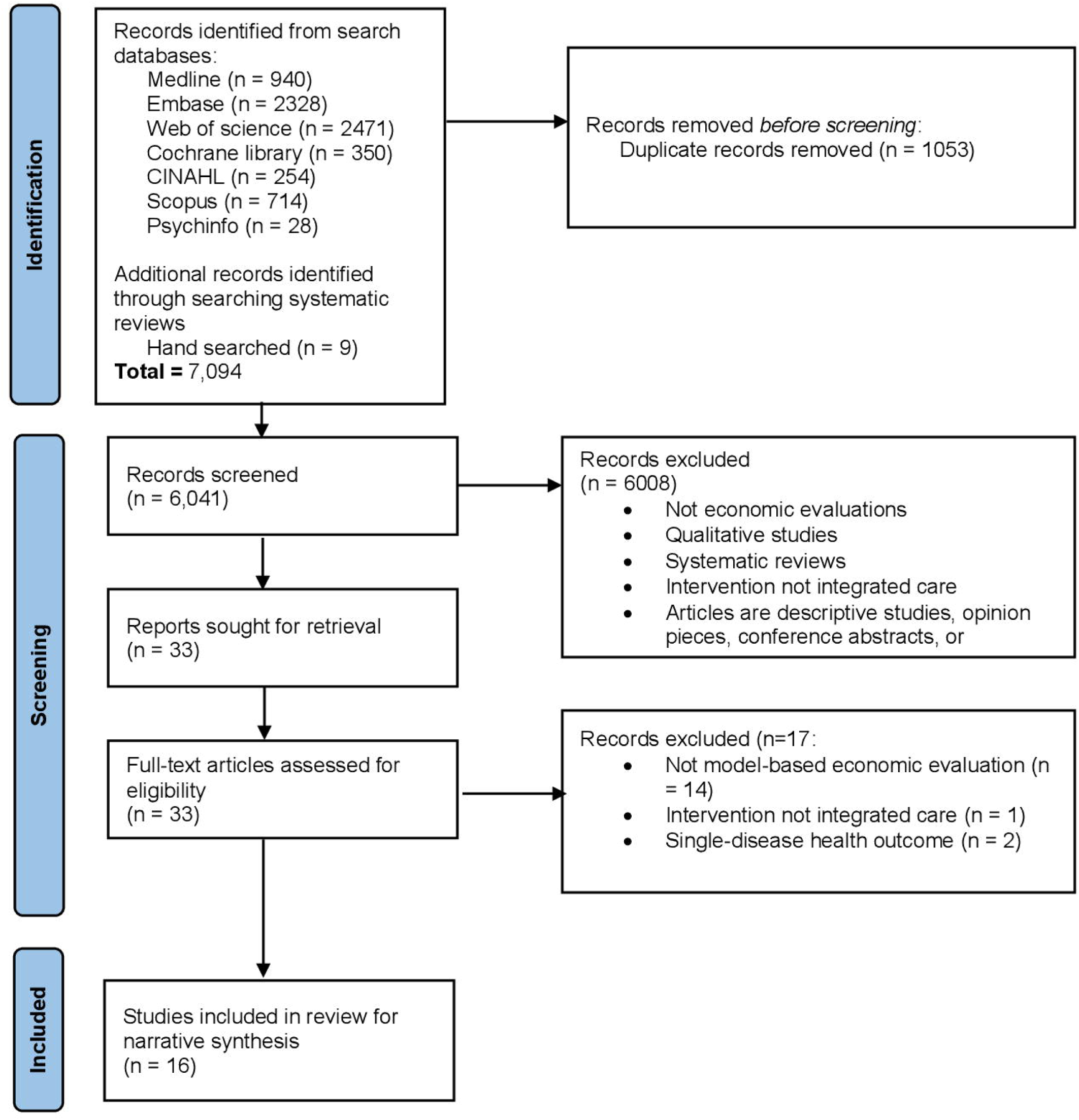
PRISMA flow chart of identification, screening, and final inclusion of articles

### Populations and interventions

#### Settings and target populations

Eleven studies were conducted in high income countries, three in lower middle-income countries (Bhutan ^62^, Jordan ^63^, and Kenya ^64^), and one in a low-income country (Uganda ^50^): see Figure 2. Modelled populations ranged between 15 and 75 years of age. In nine of the included articles, the starting population were either previously diagnosed or having hypertension ^65–67^, type 2 diabetes mellitus (T2D) ^51,52,63,68^, HIV ^50^, multiple disease risk factors ^62^, or a combination of these ^52^. Seven studies had a baseline population without disease ^64,69–74^.

**Figure 2:**
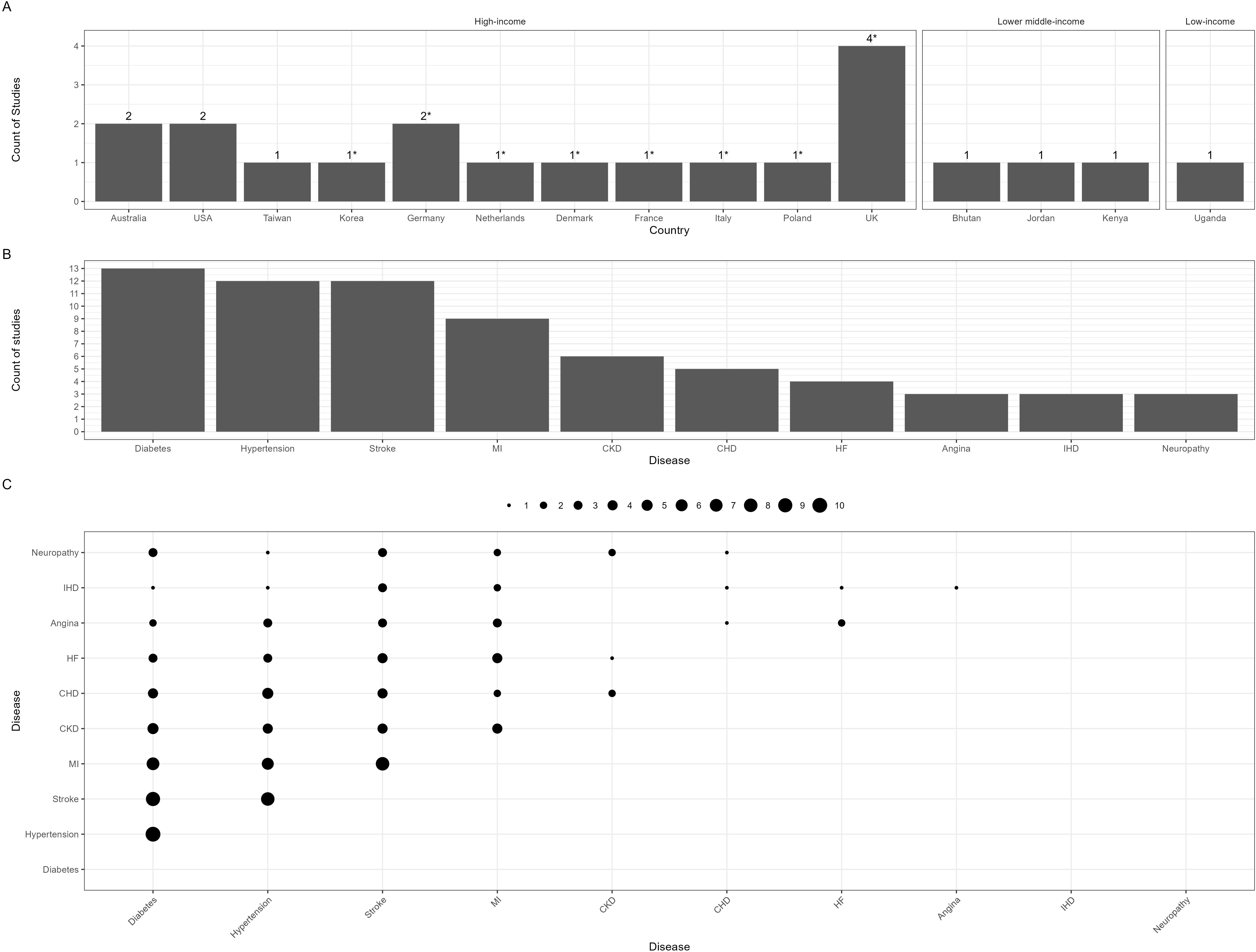
Geographical distribution and disease distribution in selected studies, Figure 2(A): Geographical distribution of included studies; USA, United States of America, UK, United Kingdom, Countries contained in one multicountry study; Figure 2(B): Top 10 most frequent diseases in selected studies, Figure 2(C): Combination of disease conditions in selected studies. Dots represent number of studies with disease combinations included.

#### Disease conditions modelled

The review focused on both concordant and discordant cardiometabolic multimorbidity. Eight of the 16 included studies had diabetes as the primary disease ^51,52,62,63,68,70,71,73^. Hypertension was the primary disease in four of the studies ^62,65,67^. Two studies in SSA had HIV as the primary disease ^50,64^. Other primary conditions included atrial fibrillation ^69^ and cancer ^74^ (Table 1). The most common pair of conditions considered was hypertension or diabetes and their related complications including stroke, myocardial infarction (MI), coronary heart disease (CHD), chronic kidney disease (CKD) (Figure 2).

**Table 1:** Characteristics of the included studies.

| Author<br>(year) | Country | Evaluation<br>type | Health<br>system<br>context | Integrated care intervention | Comparator | Main disease modelled | Other co/ multimorbidities<br>modelled |
| --- | --- | --- | --- | --- | --- | --- | --- |
| Afzali et<br>al. (2012)<br><sup>75</sup> | Australia | CUA | Primary<br>health<br>care | High level Practice nurse<br>involvement in clinic-based<br>activities | Low level Practice nurse<br>involvement in clinic-based<br>activities | Diabetes | -MI<br>-CHD (ISD)<br>-Heart failure<br>-Stroke<br>-Renal failure<br>-Blindness<br>-Amputation |
| Dukpa et<br>al. (2014)<br><sup>62</sup> | Bhutan | CEA | Primary<br>health<br>care | WHO PEN which uses an<br>integrated approach to assess<br>and manage cardiovascular risk<br>using hypertension and diabetes<br>as entry points | No screening | Hypertension and/or<br>diabetes | -Stroke<br>-CHD<br>-Retinopathy<br>-Neuropathy<br>-Nephropathy |
| Hirsch et | USA | CEA | Primary | A collaborative endocrinologist- | Usual primary care physician | Diabetes | -Hypertension |
| al. (2017)<br>52 |  |  | health<br>care | pharmacist Diabetes Intense<br>Medical Management (DIMM)<br>“Tune-Up” clinic for complex<br>diabetes patients | (PCP) care |  | -Hyperlipidemia<br>-MI<br>-CHF<br>-CHD (Coronary artery<br>disease)<br>-CKD<br>-COPD |
| Hobbs et<br>al. (2005)<br>69 | United<br>Kingdom | CEA | Primary<br>health<br>care | Different screening strategies<br>(targeted, whole-population<br>screening and opportunistic<br>screening with prompts) | No screening | Atrial fibrillation | -Ischaemic -stroke<br>-Haemorrhagic<br>stroke<br>-GI -complications |
| Howard<br>et al.<br>(2010) 72 | Australia | CUA | Primary<br>health<br>care | Primary care based screening for<br>CKD risk factors and improved<br>management | Routine glucose and blood pressure<br>control | CKD | -Hypertension<br>-Diabetes<br>-Proteinuria |
| Kasaie et<br>al. (2020)<br>64 | Kenya | CEA | Primary<br>health<br>care | A joint community outreach<br>campaign for screening and<br>treatment of HIV, hypertension, | Standard care | HIV | -Diabetes<br>-Hypertension<br>-Cardiac arrest |
|  |  |  |  | and diabetes |  |  | -MI<br>-Angina<br>-Stroke |
| Kim et al.<br>(2021) <sup>66</sup> | Republic<br>of Korea | CUA | Primary<br>health<br>care | A chronic disease management<br>program (CDMP) in patients<br>with hypertension | Usual care | Hypertension | -MI<br>-Stroke<br>-CKD<br>-Heart failure |
| Mason et<br>al. (2005)<br><sup>70</sup> | United<br>Kingdom | CUA | Primary<br>health<br>care and<br>hospital | Specialist nurse–led clinics<br>provided to<br>improve lipid and blood<br>pressure control in diabetic<br>patients | Usual care | Diabetes | -Hypertension (Part of<br>intervention)<br>-Hyperlipidemia (Part of<br>intervention)<br>-Stroke<br>-MI |
| Mousa et<br>al. (2021)<br><sup>63</sup> | Jordan | CEA | Primary<br>health<br>care and<br>hospital | Pharmacist-led care | Usual care | Diabetes | Cardiovascular disease events<br>including:<br>-Stroke<br>-CHD |
| Penaloza-Ramos et al. (2016)<br><sup>65</sup> | United Kingdom | CUA | Primary health care | Self-monitoring and self-management of blood pressure in hypertensive patients | Usual care | Hypertension | -CHD<br>-Diabetes mellitus<br>-Stroke<br>-CKD |
| Sando et al. (2020)<br><sup>50</sup> | Uganda | CEA | Primary health care | Integration of screening and treatment for hypertension, diabetes, and hypercholesterolemia into HIV care | Standard HIV care without NCD screening | HIV | -Hypertension<br>-Diabetes<br>-Hypercholesterolemia |
| Schaufli et al. (2010)<br><sup>71</sup> | Germany | CUA | Primary health care | Screening strategy for type 2 diabetes mellitus (T2D) including early detection and secondary prevention | Routine clinical practice | Diabetes | -MI<br>-Stroke<br>-Renal disease<br>-Neuropathy |
| Schouten et al. (2010)<br><sup>68</sup> | Netherlands | CUA | Primary health care | A quality improvement collaborative (QIC) focusing on patients with diabetes | Usual care | Diabetes | -MI<br>-Stroke<br>-Renal disease<br>-Neuropathy |
| Schuetz et al. (2013) <sup>73</sup> | Denmark, France, Germany, Italy, Poland, and the United Kingdom | CUA | Primary health care | A vascular disease Health Checks program which integrates the prevention, early detection, and treatment of type 2 diabetes, hypertension, dyslipidemia, and smoking. | Usual care | Diabetes | -Coronary artery disease<br>-CHF<br>-Stroke<br>-Hypertension<br>-Obesity<br>-Metabolic syndrome<br>-Cancers (breast, lung, colon) |
| Schultz et al. (2021) <sup>67</sup> | USA | CUA | Primary health care | A pharmacist-led medication therapy management (MTM) clinic | Usual care | Hypertension | - MI<br>-Stroke<br>-Other CVD events (peripheral artery disease, angina, and transient ischemic attack)<br>-Chronic CVD<br>Tunnel states: -stroke recovery<br>-MI recovery |
|  |  |  |  |  |  |  | -Other recovery |
| Wang et al. (2006)<br>74 | Taiwan | CEA | Primary care and hospital | A community-based Integrated Screening (KCIS) programme for chronic diseases | No screening | Cancers: colorectal, breast, cervical and oral cancer | -Type 2 diabetes<br>-Hypertension |

CEA, cost effectiveness analysis; CUA, cost utility analysis; CKD, chronic kidney disease; CVD, cardiovascular disease; MI, myocardial infarction; GI, gastrointestinal; CHD, coronary heart disease; CHF, congestive heart failure; IHD, Ischaemic heart disease; PEN, Package of essential NCD interventions

#### Interventions modelled

The integrated care models or interventions evaluated varied from health care provider-led interventions (e.g., pharmacist-led), integrated screening and treatment, comprehensive disease management programs, and a quality improvement intervention. The details and characterisation of the integrated care interventions are presented in Table 1 and online supplemental appendix table S3, respectively. There was a high level of diversity in the interventions modelled under the definition of integrated care in the selected studies. Two studies conducted in Jordan ^63^ and USA ^67^ evaluated pharmacist-led care and medication therapy management (MTM) for diabetes and hypertension respectively. Hirsch et al.,^52^ evaluated collaborative endocrinologist-pharmacist intense medication management for diabetes in the USA. One study in Australia ^51^ evaluated high level patient nurse involvement, while another study in the UK ^70^ focused on specialist nurse-led clinics for diabetes. Integrated screening and treatment constituted most of the included studies. The study conducted in Bhutan ^62^ and a multicountry study in six European countries ^73^ evaluated integrated screening and treatment and management for diabetes, hypertension among other related risk factors. A study in Uganda ^50^ evaluated integrated screening and treatment of NCDs into HIV care. Early detection and secondary prevention of diabetes was evaluated in one study in Germany ^71^ while a study in Australia ^72^ evaluated primary care-based screening for chronic kidney disease risk factors and improved management. Community-based integrated screening was evaluated in two studies in Kenya ^64^ and Taiwan ^74^. One UK study examined whole population screening and opportunistic screening compared to usual care ^69^. Self-monitoring and management of blood pressure in hypertensive patients was evaluated in one UK study ^65^ while another study conducted in the Netherlands ^68^ evaluated a quality improvement collaborative for patients with T2D.

### Analytic approaches used

#### Economic approach

Cost utility analysis (CUA) was used by 13 studies ^50,52,62,64–68,70–73,75^, while three studies ^63,69,74^ were cost-effectiveness analysis studies whose outcomes were not based on utilities (Table 1).

#### Discount rates

Discount rates used in the selected studies varied among countries due to the conventions used in different countries. The discount rates for costs ranged between 3% and 5% while the discount rates for outcomes ranged between 1.5% and 5% (Table 2).

**Table 2:** Characteristics of the decision analytic models developed in the economic evaluations.

| **Author**<br>**(year)** | **Model type** | **Time horizon** | **Perspective** | **Discount rate** | **Costs and resource use parameters included** | **Disease parameters included** | **Analysis of sensitivity and uncertainty** | **Existing model adapted** |
| --- | --- | --- | --- | --- | --- | --- | --- | --- |
| Afzali et al. (2012) | SMM | 40 years | Health care system | 5% | Patient-level data on hospital and primary care services from 70,340 patients with a mean duration of 4.5 years of follow-up. | History of diabetes<br>History of comorbidities (e.g., peripheral vascular disease [PVD]) History of diabetes-related complications. Time-varying risk factors (HbA1c, BP, TC, HDL, smoking status) | PSA (non- parametric bootstrapping)<br>First-order uncertainty (i.e., variability) minimised by performing 1,000 repeated Simulations per patient in the model. | No |
| Dukpa et al. (2014) | Decision tree and a SMM | Lifetime | Societal | 3% | Screening<br>BP monitoring<br>Treatment and follow up of diabetes<br>Treatment and follow up of hypertension | Prevalence, proportions of hypertension in diabetes patients<br>Transitional probabilities - developing complications or death<br>Sensitivity, and specificity of screening tools | PSA for parametric uncertainty<br>First-order uncertainty (i.e., variability) was minimised by performing 1,000 repeated Simulations per patient in the model. | No |
| Hirsch et al. (2017) | Individual sampling model | 2 years, 5 years, and 10 years | Health care system and payer | 3% | Intervention costs: Clinical pharmacists (including follow-up phone contact, nonpatient administrative cost, and patient no-show time, and recognizing non-scheduled weeks for the DIMM clinic) Inpatient, outpatient, ambulatory, treatment, and other costs. | Incidence, risk reduction of diabetes and related complications BMI, weight, blood pressure, high-density lipoprotein cholesterol, low-density lipoprotein cholesterol, total cholesterol, triglycerides, fasting plasma glucose, A1c, and glomerular filtration rate. | 1-way and 2-way, and PSA were conducted using upper and lower limits of 95% confidence intervals (CIs) for clinical outcome parameters Probabilistic analyses simulated 1,000 replications | Archimedes model |
| Hobbs et al. (2005) | DES | Lifetime | Health system and patient perspective | 3.5% | Screening costs<br>Costs of stroke<br>Costs of GI bleed<br>Cost of Warfarin treatment | Prevalence & incidence of AF, first ischaemic stroke, GI bleed<br>Rate, risks and probabilities of AF, stroke<br>Relative risk, and risk reduction with medication. | PSA was undertaken using distributions for variables where variation around the point estimates used in the model was available. For each patient group and screening type, plus no screening | No |
|  |  |  |  |  |  |  | A total of 10,000 replications was performed, with summary data for each replication representing 10,000 patients. |  |
| Howard et al. (2010) | SMM | Lifetime | Health care funder perspective | 5% | Drug costs<br>Out-patient health care utilisation including consultation costs for different providers e.g., GP, dietician, podiatrist, nephrologist, diagnostic tests | Relative risks of CVD death<br>CVD events<br>Progression to microalbuminuria<br>Progression to end-stage kidney disease (ESKD) for hypertension and diabetes patients | PSA was conducted | No |
| Kasaie et al. (2020) | Individual sampling model | 15 years | National and regional | 3% | Costs of standard care<br>Costs of screening for hypertension and diabetes<br>Costs of acute care for cardiac arrest, MI, angina, stroke, post-CHD, post-stroke<br>Costs of HIV, hypertension, | CVD natural history<br>CVD Risk - 10-year risk categories<br>Probability of first CHD event (cardiac arrest, MI, Angina)<br>Acute and annual Mortality following CVD event | One-way sensitivity analysis was performed by varying the value of selected parameters to +/-15% of the original values | No |
|  |  |  |  |  | and diabetes treatment | NCD treatment effectiveness<br>HIV, Hypertension, and diabetes prevalence<br>ART coverage<br>HIV incidence |  |  |
| Kim et al. (2021) | SMM | Up to 60 years | Health care payer | 5% | Direct medical costs for hypertension, MI, stroke, CKD, heart failure<br>Additional costs including pay-for-performance incentives to patients<br>Costs for health support services | Incidence rates<br>Mortality rates<br>Effects of CDMP - Hazard ratios | Sensitivity analysis conducted based on probability, utility, and cost (Probabilities varied based on their 95% confidence intervals (CIs), utilities in between a $\pm 20\%$ range, and costs up to 92.0 USD (100 000 KRW)).<br>A sensitivity analysis at a range of around 20% excluding the cost of the policy was conducted. | No |
| Mason et al. (2005) | SMM | Lifetime | Health care system | 5% | Costs of drugs, acute and chronic treatment after stroke and MI | Risk of suffering a stroke or MI based on age, sex, and cardiovascular risk factors | Uncertainty surrounding estimates explored using CEACs, generated from Monte Carlo analyses making | No |
|  |  |  |  |  |  | Risk of death for patients who have had a stroke or MI, adjusted by age and sex<br>Mortality adjusted for diabetes<br>Mortality from non-cardiovascular causes<br>Risk ratios for blood pressure and lipid lowering | 10,000 evaluations of each model.<br>Parameters were sampled randomly from distributions for costs of acute and chronic stroke and MI, and risk ratios for BP and lipid lowering. |  |
| Mousa et al. (2021) | SMM | 10-years | Public health provider | 4.75% | Medical procedure costs for CVD events<br>Outpatient and inpatient costs<br>Medication for outpatient visit<br>Utilisation of medication | Changes in BP, HA1c, total cholesterol, HDL-cholesterol<br>Risk of T2D patients experiencing CVD events over 10 years | PSA was conducted for the base-case analysis to allow all variables to vary simultaneously by assigning a probability distribution for CVD risk, LYG, and cost, where gamma distribution was assigned for cost data, beta distribution for 1- to 10-year CVD risk, and Lognormal distribution for the relative risk of stroke recurrence following primary | No |
|  |  |  |  |  |  |  | CHD and stroke.<br>Monto-Carlo simulation with 10,000 iterations was performed to construct a CEAC. |  |
| Penaloz<br>a-Ramos<br>et al.<br>(2016) | SMM | Horizon<br>varied<br>from 30<br>years<br>(lifetim<br>e) to<br>between<br>1 - 20 | UK NHS<br>and<br>Personal<br>social<br>services | 3.5% | Costs for ongoing BP monitoring in primary care, self-management, and prescription of antihypertensive agents<br>Intervention: Self-management, equipment, and training costs. Replacement costs for the equipment and training were included at five yearly intervals<br>Costs for acute and chronic cardiovascular events | Blood pressure control<br>Stroke risk<br>Cardiovascular risk<br>Mortality. | Data were entered into the model as distributions in order that a PSA could be undertaken to incorporate parameter uncertainty<br>Gamma distributions were fitted to all costs obtained from the TASMIN-SR trial and beta distributions were applied to the utility values.<br>The PSA was run with 10,000 second-order Monte Carlo simulations and cost-effectiveness planes and cost-effectiveness acceptability curves constructed to estimate the probability of self- management being cost-effective at different willingness-to-pay thresholds. | No |
| Sando et al. (2020) | Epidemiologic-cost model | 10 years | Health care system | 3% | Costs of medical consultation, lab, and medicines for hypertension, T2DM, and high cholesterol patients)<br>Cost of hospitalisation for fatal or non-fatal CVD event and treatment costs | NCD risk factors<br>Prevalence of HIV among 15–49-year-olds<br>Proportion of HIV-infected enrolled in ART programs<br>Prevalence of hypertension, DM, hypercholesterolemia<br>Proportion on medication for hypertension, DM, and high cholesterol<br>Treatment efficacy for NCD risk factors (RR of stroke and coronary events for hypertensive, diabetic, and high cholesterol patients) | Univariate sensitivity analyses, where varied one input parameter at a time independently while maintaining values for the other input parameters unchanged. | No |
| Schaufli et al. (2010) | SPLMM | Lifetime | German statutory health insurance | Costs discounted at 5% QALYs not discounted | Costs for interventions: Screening test verification of diagnosis costs Prevention using drugs and lifestyle interventions Costs for complications | Age-dependent prevalence of pre-diabetes and T2D Incidence of pre-diabetes and T2D Incidence rates for complications Mortality rates Risk reductions due to prevention | Varying the following parameters within reasonable limits: Costs of early detection and prevention. Discounting rates for costs and utilities, Participation rates in the target population (5%, 15%, 45%, 30%), effectiveness of early detection and prevention. | No |
| Schouten et al. (2010) | SMM | Lifetime | Health care system | Costs: 4.50 % Effect sizes: 1.50% | Intervention costs (program-management costs of the Dutch Institute of Health care Improvement Costs of the participating health care providers (time and related costs) spent on the project, local overhead for each site, per patient | Life expectancy (Patient ageing) HRQoL % of nonsmokers % of patients with HbA1c <7 and <8.5% Hazard ratios for excess cardiovascular morbidity and mortality associated with diabetes | One-way sensitivity analyses were conducted to test the robustness of the model results. PSA was performed to estimate the uncertainty surrounding the ICER. | No |
|  |  |  |  |  | annually).<br>Health care costs | Age-related risk of diabetes-specific complications |  |  |
| Schuetz et al. (2013) | Individual sampling model | 30 years (Lifetime) | Health care system | 3% | Costs associated with the health check, subsequent testing and treatment, and cardiovascular events. | Incidence and prevalence of diagnoses of T2D, major adverse cardiovascular events (MACE, first occurrence of myocardial infarction, stroke, or cardiovascular death), and a composite of serious microvascular complications (first occurrence of diabetes related blindness, CKD, end stage renal disease, renal death, foot ulcer or foot amputation)<br>Medication use (e.g., anti-hypertensive, statin, and anti-diabetic treatments)<br>Disease burden in the country (including type 2 diabetes, nephropathy, retinopathy, neuropathy, myocardial infarction, stroke, cardiovascular death, and mortality). | Not clearly reported | Archimedes model |
| Schultz et al. (2021) | CTM<br>C<br>Model | 10 years | Payer perspective | 3% | Costs of patient visits to MTM clinic<br>Costs for primary prevention<br>Costs of CVD events (states in the model including Stroke-tunnel, MI-tunnel, and other-tunnel states) | Mean changes in A1c, blood pressure, emergency department and hospital admissions<br>10-year CVD risk<br>Transition probabilities (Same CVD event, primary prevention to CVD event, recovery state of one CVD event to another CVD event)<br>Mortality risk | One-way and PSA were performed. Risk ratios were varied by 10% above and below the base-case values, utilities by 20%, and costs by 50% to 200%.<br>A 10 000-iteration Monte Carlo simulation was used to perform the PSA. CEACs were generated to display the Monte Carlo simulation results. | No |
| Wang et al. (2006) | SMM | 20 years or 99 | Health care system | 5% | Screening costs<br>Manpower cost<br>Confirmation costs | Disease progression<br>Transition probabilities<br>Natural history for each disease | Not reported | No |
|  |  | years<br>(Lifetime) |  |  | Average costs for terminal care | risk factor prevalence<br>Survival<br>Complication rates e.g.,<br>perforation in colonoscopy<br>Association between non-malignant chronic diseases and cancers<br>Screening performance parameters: Sensitivity and specificity |  |  |

CTMC, Continuous time Markov chain; DES Discrete event simulation; PSA Probabilistic sensitivity analysis; SMM, Simulated Markov Model; SPLMM, Simulated patient-level Markov model

#### Model type

Of the 16 selected economic evaluations, nine used simulated Markov models to evaluate the impact of the integrated care interventions ^51,63,65,66,68,70,71,74^. Of these studies, eight studies ^51,62,63,66,67,70–72^ had an annual cycle length while one study in the UK ^65^ used a 6-month cycle length. Three studies used individual sampling models (microsimulation models) which incorporated the patients’ unique medical histories and characteristics and complexities of the health system and multimorbidity in the modelling ^52,64,73^. The study conducted in Uganda used an epidemiologic-cost model to estimate costs and effects of integrated screening and treatment ^50^. One study conducted in the USA used a discrete event simulation (DES) for their evaluation from the health system and payer perspective ^69^. One study conducted in the USA used a semi-Markov model in the evaluation to incorporate time-varying mortality ^67^.

#### Model perspective

Seven of the included studies performed the economic evaluations from a health care system perspective ^50,51,63,64,68,70,73,74^ while only three studies included a societal or patient perspective in their evaluations ^62,65,69^. Three studies performed their evaluations from both health system and payer perspectives ^52,65,69^. One study used a health care funder perspective ^72^ and another study in Germany was performed from the perspective of the German statutory health insurance ^71^.

#### Model horizon

The time horizons in the included studies ranged between two years and a lifetime horizon. Ten of the selected studies used a lifetime horizon ^62,65,66,68–74^. Four studies conducted in the USA, Uganda, and Bhutan used a 10-year horizon ^50,52,63,67^, while one study in Kenya used a 15-year horizon ^64^. In addition to a lifetime horizon, three studies in Taiwan, UK, and USA simulated between one and 20 years horizon ^52,65,74^.

#### Model adaptations

Eight of the selected studies either adapted existing models or used them to generate inputs. One study conducted in Australia ^51^ used the UK Prospective Diabetes Study (UKPDS) outcomes model to estimate costs and effects of the alternative models of care ^76^. Studies conducted in Bhutan (Mousa) and the Netherlands ^68^ used the UKPDS risk engine to calculate CVD risk estimates for their models ^77,78^. A study conducted in the USA ^52^ and another in six European countries and the UK ^73^, used the Archimedes model to estimate costs and QALYs. The Archimedes model is validated and has been widely used to model diabetes ^79,80^. The study in Uganda ^50^ used the Globorisk model to obtain estimates of CVD risk, while one study in the USA ^67^ used Framingham risk equations ^81^. The study conducted in Kenya used an existing population-based model of HIV dynamics (SPECTRUM) combined with a microsimulation ^64^.

#### Quality assessment

The mean quality score in the included studies was 81% [57% - 93%] (Table 3). The selected studies showed good quality of reporting on the model structure domain (89%). The studies showed adequate reporting and justification of the decision problem and objective, perspective and scope of the model, rationale for the structure, assumptions, model type, horizon, cycle length and disease pathways. However, only four studies provided a clear statement on whether all feasible and practical options were evaluated ^50,62,71,74^ while it was not clear in 11 studies. Despite almost 90% of studies stating the model perspective, the primary decision-maker for the analysis was not clear in almost half of the selected studies. Majority of studies adequately reported costs, utilities and their sources, data identification, and methods of incorporating data (76%). Notably, patient indirect costs and productivity losses were not reported for most studies, including those that used a societal perspective. Furthermore, only one study evaluating a quality improvement collaborative reported program implementation costs such as project management and local overhead costs for participating facilities ^68^. However, deficiencies were noted in the assessment of uncertainty where only two studies reported having used all the four principal types of uncertainty with majority not justifying omission ^65,69^. Only five out of the 16 selected studies performed subgroup analysis by disaggregating the final cost-effectiveness results by gender or age group ^50,64,65,69,71^. More than 80% of the studies adequately reported and addressed structural and parameter uncertainty while less than half assessed methodological uncertainty or heterogeneity by running the models separately for different subgroups. For the consistency domain, higher scores were achieved for external consistency as compared to internal consistency. Only five studies justified the internal validity of their model ^51,52,68,69,73^.

**Table 3:**
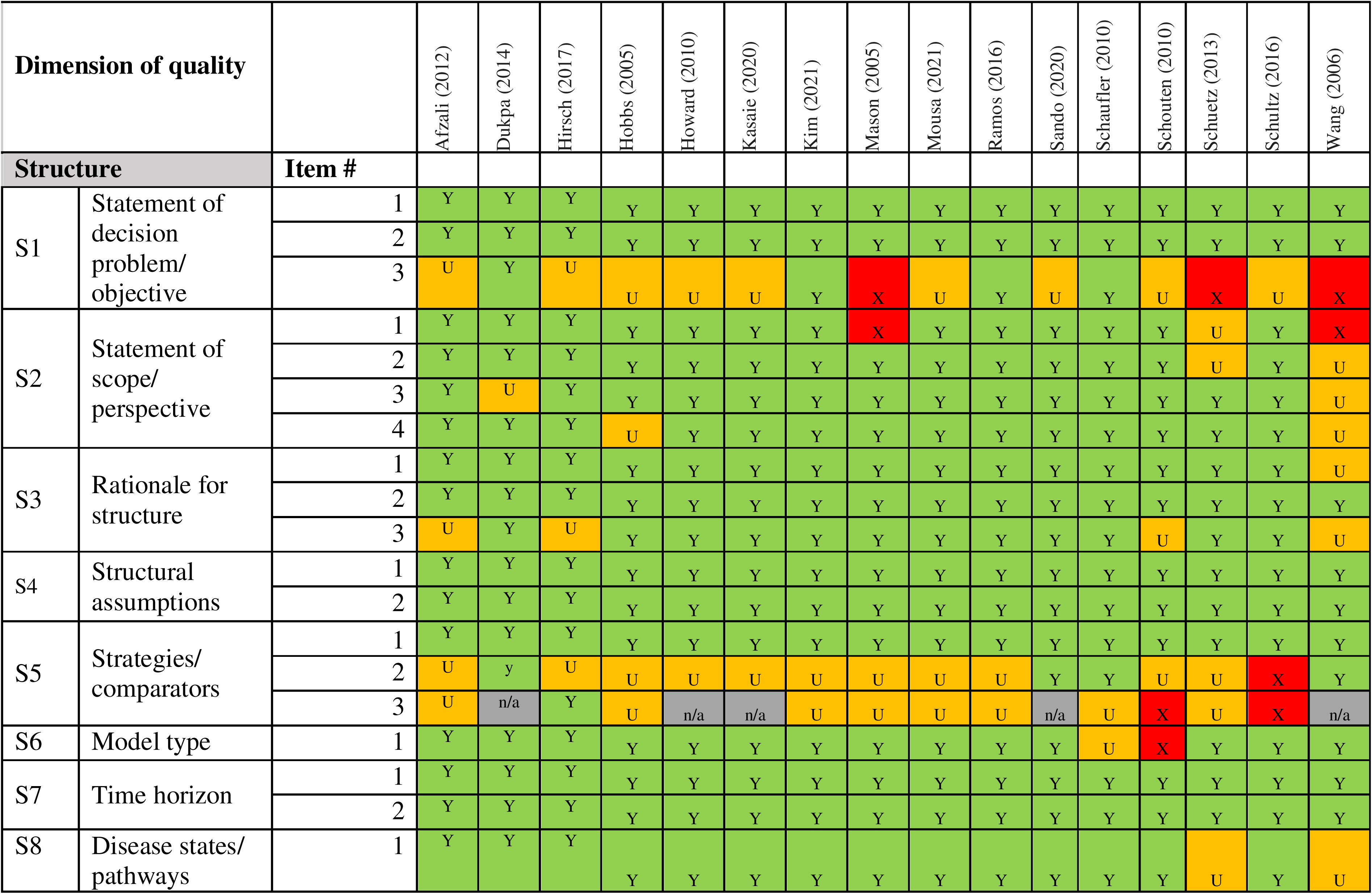
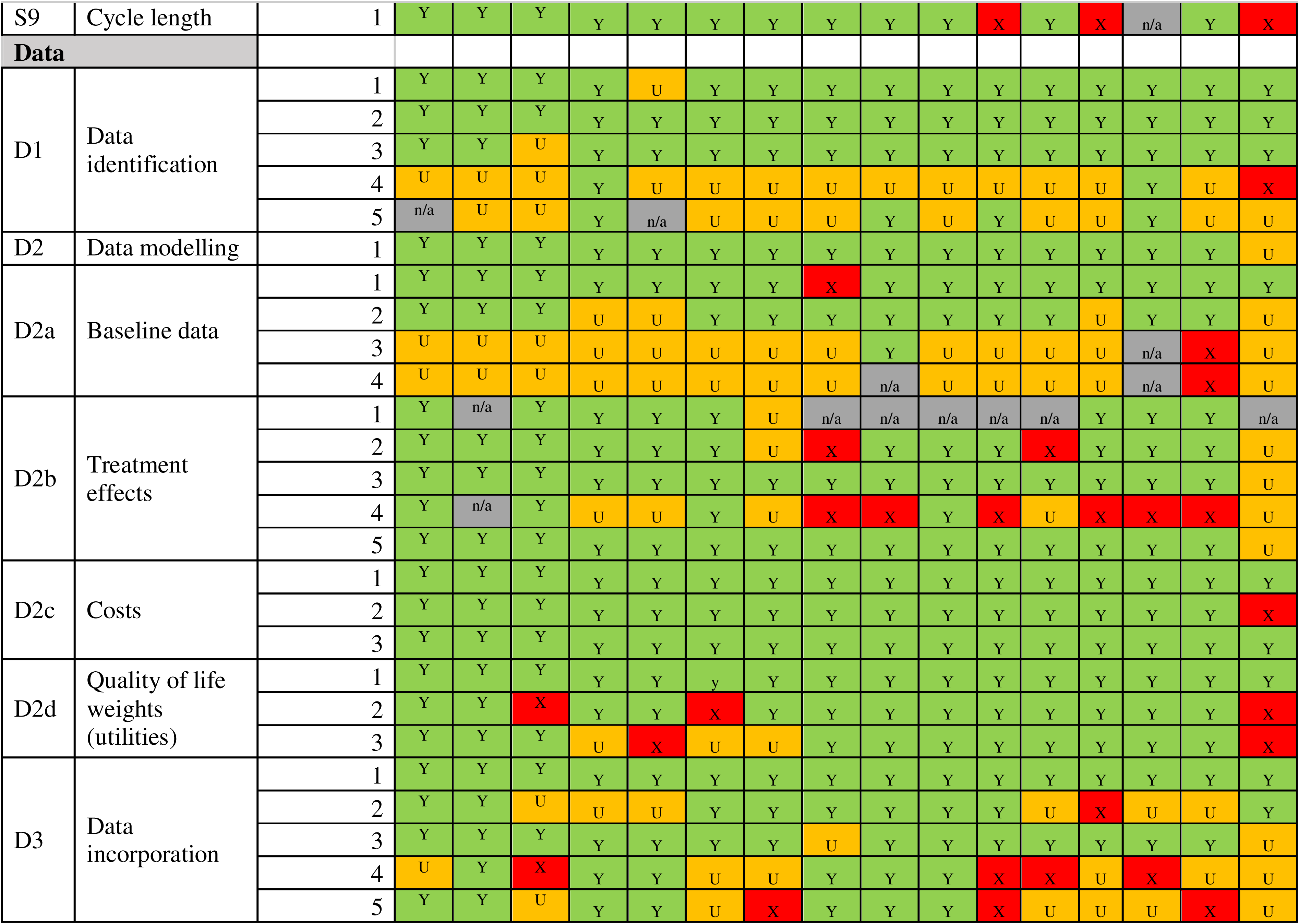
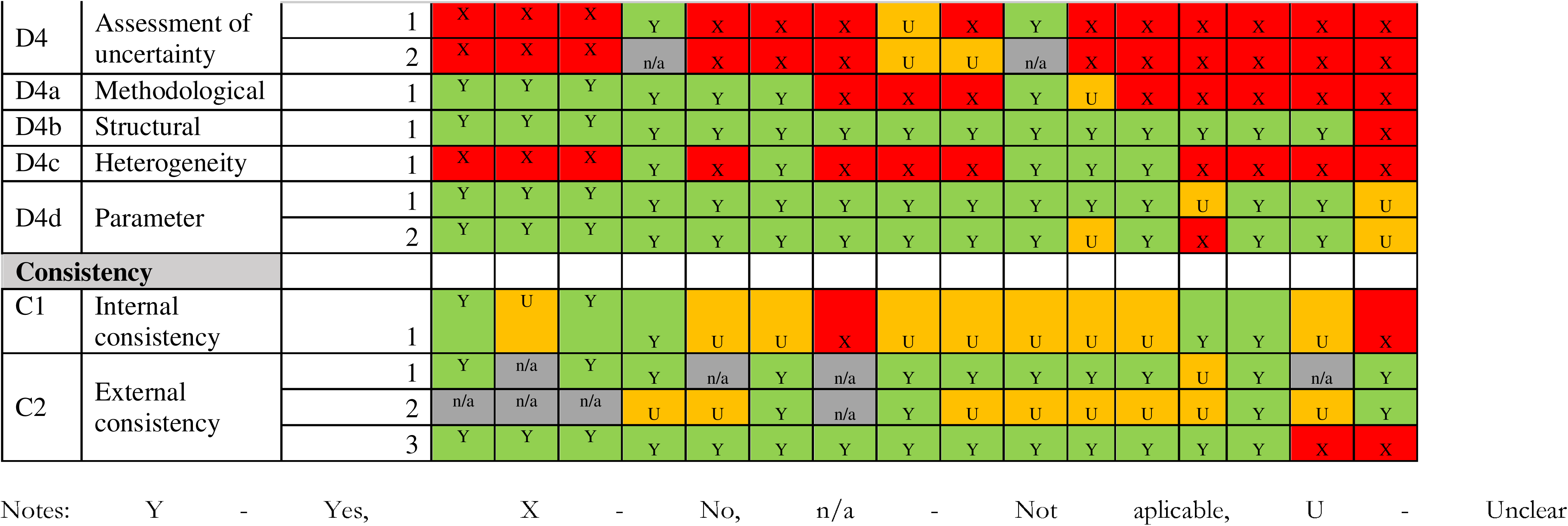
Quality assessment of included studies.

## Discussion

A total of 16 studies were included in this systematic review, majority of which were conducted in high-income countries than LMICs. There was significant heterogeneity in the types of integrated care models evaluated as well as the chronic disease focus in the included studies. However, most of the studies used simulated Markov models and focused on hypertension or diabetes as the primary disease condition. While the health outcome metrics reported were consistent, there was poor reporting of data validation approaches against local data, the quality of data incorporated in the models, and internal and external consistency. Furthermore, individual patient simulations were better able to capture complexities of integrated care interventions and multimorbidity.

The majority of the integrated care models in the included studies were integrated screening and treatment interventions at health facility and community level. This is consistent with existing literature on integrated care for multiple chronic diseases focused on integrating screening and treatment ^82–85^. The primary disease conditions in the evaluations were diabetes or hypertension which supports literature from existing studies on integrated care ^86–89^. The only two studies from this review focused on infectious disease and NCD integration were conducted in SSA countries. This corroborates studies on integrated care in SSA which have focused on HIV and NCD integration in the region ^89–92^. A plausible reason for this integration is the heightened risk of NCDs among PLHIV who have higher life expectancy due to improved therapies and management structures. However, the findings indicate the need to evaluate models of integration for concordant cardiometabolic multimorbidity occasioned by the rising burden of these conditions ^24^.

The majority of the studies performed the model-based economic evaluations from a health system perspective ^50,51,63,64,68,70,73,74^, while only three studies included a societal or patient perspective in their evaluation ^62,65,69^. Of note is that the studies that included a patient perspective did not report including the indirect costs such as informal care and productivity losses in addition to the direct patient costs as recommended by existing health economic guidelines ^93^. Most of the selected studies did not provide implementation costs of the integrated care models. In fact, only one study that evaluated a quality improvement collaborative included the program implementation costs, such as program management costs and local overhead costs for each participating facility, as part of their inputs for the economic model ^68^. Guidelines for economic evaluations recommend a lifetime horizon for interventions that impact costs and outcomes over a patient’s lifetime, with shorter horizons being appropriately justified ^93^. However, a third of the selected studies did not include a lifetime horizon and studies that used 10-year ^50,52,63,67^ and 15-year ^64^ horizons did not provide clear justifications. Due to the long-term effects of multiple chronic conditions hence the need to seek continuous care, a lifetime horizon is appropriate in economic evaluations of integrated care.

Majority of the DAMs in the selected studies were Markov models. The evaluation of a chronic disease programme in Korea used a Markov model that assumed identical progression probabilities after development of complications in the two groups, which may underestimate or overestimate the outcomes ^66^. Similarly, a Markov model to evaluate the WHO PEN model for hypertension and diabetes in Bhutan assumed similar treatment outcomes for comorbidity and diabetes alone ^62^, yet there may be considerable variation in the disease outcomes for a single disease compared to a patient with comorbidity. Markov models are considered limited in assessing complex health care interventions due to their limitations in capturing individual dynamics ^48,94,95^. In the case of integrated care for multiple chronic diseases, the multiple disease states that would exist with multimorbidity and complex interactions within an integrated health system, such as repeated interactions with health care, may not be adequately represented. Furthermore, assumptions of constant transition probabilities, as was observed with majority of the selected studies using Markov models, overlook potential temporal changes and relationships that may exist, and important interplays between health system elements and patient behaviours may be a potential limitation. For such interventions, more advanced methodologies such as individual sampling models, dynamic simulation, or system dynamics models are recommended to better encompass the complexities ^48,96^. For instance, one of the included studies conducted in Kenya ^64^ used a microsimulation model and was able to model CVD dynamics at individual level by modelling impact on patients based on their levels of CVD risk, which was not captured by the cohort level models.

Despite most of the included DAMs scoring well in their quality of reporting, there was poor reporting with regards to accounting for uncertainty and validation in the studies. Philips et al., (2006) recommend distinguishing between the four principal types of uncertainty when reporting economic evaluations i.e. methodological, structural, heterogeneity, and parameter uncertainty ^61^. Similar to our findings, other reviews on economic evaluations show that economic evaluations have scored highly on reporting structural and parameter uncertainty through one-way, two-way, and probabilistic sensitivity analyses, and scoring poorly on reporting heterogeneity and methodological uncertainty ^97,98^. With regards to heterogeneity, only about a third of the selected articles reported performing subgroup analysis. For a health care intervention such as integrated care which impacts the general population, it is important to assess its differential impact on different groups to account for the equity considerations in case the interventions are to be scaled up, an important consideration for policy makers.

The findings of our systematic review have important implications for health care decision making for chronic disease prevention and control in LMICs where the burden of multimorbidity is increasing ^24,27^. Given the demonstrated potential benefits of integrated care for patients with chronic disease multimorbidity in other settings, the scarcity of economic evaluations using DAMs in LMICs signals a substantial data gap that limits evidence-based decision making for chronic disease prevention and control. Therefore, there is an urgent need for more economic evaluations using DAMs to inform integrated care models of healthcare delivery that are tailored to different contexts in LMICs for cardiometabolic multimorbidity and other common multimorbidity patterns. Considering the social and health inequalities and disparities in healthcare provision that have been evidenced in LMICs^99,100^, health economists and analysts should consider the use of individual sampling models that will capture the complexity of different integrated care configurations and patients interactions with the health care models in the context of multiple diseases using a lifetime horizon. This ensures that recommended integrated care models are deemed cost-effective, context-specific, and sustainable.

This systematic review is the first to synthesise evidence on model-based economic evaluations conducted to evaluate integrated care for cardiometabolic multimorbidity. The key strength of our study is the broad review of the literature that was designed to capture studies with a wide heterogeneity, and synthesized the findings to provide more understanding of how integrated care can been modelled using DAMs. Our study has some limitations. Firstly, we only included original articles published in the English language, hence there is a chance that relevant studies published in other languages may have inadvertently been excluded from the review. Furthermore, non-inclusion of grey literature such as thesis, reports or guidance documents may have left out some relevant studies that meet the inclusion criteria. Despite these limitations, our systematic review contributes to the existing literature by synthesizing and appraising the current evidence base on the use of DAMs to model the economic impact of integrated care interventions for cardiometabolic multimorbidity.

## Conclusion

Model-based economic evaluations (including simulated Markov models, individual sampling models, discrete event simulation, and epidemiologic cost models) have been used to evaluate integrated care interventions for cardiometabolic multimorbidity, most often considering hypertension and/ or diabetes in high-income countries. Future studies could improve in their consideration of uncertainty and validation and should consider methods such as microsimulation that can more easily describe repeated interactions with health care. More studies should consider inclusion of patient costs, which are a domain of potential benefit from integrated care. More studies using DAMs to evaluate integrated care for cardiometabolic multimorbidity are needed to inform decision making in LMICs, especially individual sampling models that capture the complexity of integrated care delivery within specific contexts for different populations with multimorbidity.

## Supporting information

Supplemental appendix Section S1

Supplemental appendix Section S2

Supplemental appendix Section S3

## Data availability statement

Data sharing not applicable as no datasets generated and/or analysed for this study. Not applicable.

## Ethics statements

### Patient consent for publication

Not applicable.

## Ethics approval

This study does not involve human participants.

## Acknowledgements

The authors would like to thank Ms. Louise Falzon of Sheffield Centre for Health and Related Research (SCHARR), School of Medicine and Population Health, University of Sheffield for help in the development of the search strategy, and the Wellcome Trust for funding the study.

## Contributors

EW is the study guarantor; EW, PD, DG, and RA conceptualized the study; EW, PD, DG, and RA designed the study; EW, JO, CA, and PO performed the literature search; EW performed the analysis; EW and PD developed the visualizations; EW drafted the first version of the paper; all authors provided critical review of the draft and suggestions for revision.

## Funding

EW acknowledges funding support from the Wellcome Trust as part of a doctoral training grant [218462/Z/19/Z] at the University of Sheffield.

## Competing interests

None declared.

## Provenance and peer review

Not commissioned; externally peer reviewed.

## Supplemental material

This content has been supplied by the author(s). It has not been vetted by BMJ Publishing Group Limited (BMJ) and may not have been peer-reviewed. Any opinions or recommendations discussed are solely those of the author(s) and are not endorsed by BMJ. BMJ disclaims all liability and responsibility arising from any reliance placed on the content. Where the content includes any translated material, BMJ does not warrant the accuracy and reliability of the translations (including but not limited to local regulations, clinical guidelines, terminology, drug names and drug dosages), and is not responsible for any error and/or omissions arising from translation and adaptation or otherwise.

## Notes

### Competing Interest Statement

The authors have declared no competing interest.

