## Supplemental appendix table S1 for "Application of decision-analytic models to inform integrated care interventions for cardiometabolic multimorbidity: A systematic review"

**Table S1: Definition of terms**

| **Terms** | **Definition** |
| --- | --- |
| **Cost-effectiveness studies** | A form of comparative economic analysis that evaluated two or more alternatives in terms of relative cost and outcomes, where the outcomes are measured in a single natural unit (e.g., life years gained, disease case averted, new cases detected) |
| **Cost-utility studies** | A type of cost-effectiveness analysis where the outcomes are expressed by a generic measure of health status that considers both the effect on mortality and morbidity e.g. quality-adjusted life years (QALYs) and disability-adjusted life years (DALYs)) |
| **Cardiometabolic multimorbidity** | The existence of two or more chronic diseases in the same individual, at least one of which was a cardiometabolic disease including cardiovascular diseases (e.g., coronary heart disease, cerebrovascular disease, peripheral arterial disease, rheumatic heart disease, congenital heart disease, and deep vein thrombosis), metabolic syndrome, diabetes mellitus, and hypertension in the same individual. |
| **Concordant multimorbidity** | The co-existence of two or more chronic diseases all of which are cardiometabolic diseases. i.e., are pathophysiologically related and have similar management approaches and treatment plans (e.g., type 2 diabetes and hypertension) |
| **Discordant multimorbidity** | The existence of two or more chronic diseases at least one of which is a cardiometabolic disease i.e., pathophysiologically unrelated and require different management approaches (for example type 2 diabetes and HIV). |
| **Integrated care** | Health service delivery containing two or more components of the chronic care model (CCM), as defined by Wagner (64,65), and in line with previous research for the management of cardiometabolic multimorbidity, and at least one element of Singer et al., (2011) (66) framework for measuring integrated patient care for patients with multiple or complex chronic conditions. |
