## Supplemental appendix table S2 for "Application of decision-analytic models to inform integrated care interventions for cardiometabolic multimorbidity: A systematic review"

**Table S2: Inclusion and exclusion criteria according to the PICO framework**

|  | **Inclusion** | **Exclusion** |
| --- | --- | --- |
| **Population** | Adults (18 years and above) at risk of or having either concordant or discordant cardiometabolic multimorbidity | Individuals below 18 years, adolescents, and children. |
| **Intervention** | Integrated care models or interventions in health care delivery as per the study definitions | Interventions not considered integrated care as per the study definition |
| **Comparator** | Alternative integrated care models or interventions  Usual care for patients with chronic diseases |  |
| **Outcomes** | Cardiometabolic multimorbidity as per the study definition | Single-disease focused studies and other types of multimorbidity that do not include at least one cardiometabolic disease. |
