## Supplemental appendix table S3 for "Application of decision-analytic models to inform integrated care interventions for cardiometabolic multimorbidity: A systematic review"

**Table S3: Components of the integrated care models in the included studies**

| **Author (year)** | **Community resources and policies** | **Health care organization** | **Self-management support** | **Delivery system design** | **Decision Support** | **Clinical information System** | **Coordinated care (between and within care teams)**  **Continuity of care**  **Patient-centred care** |
| --- | --- | --- | --- | --- | --- | --- | --- |
| Afzali et al. (2012) |  | Organisation supports PN involvement in clinic-based activities | Provision of self-management services by practice nurses | Support for non-physician personnel (PN) involvement in self-management support and continuous monitoring of patients and the practice. |  |  | PN involvement in clinic-based activities (coordinated care)  Monitoring of progress (continuity of care) |
| Dukpa et al. (2014) |  | Organisations support the implementation of PEN interventions | Provision of education to promote self-management | Training of non-physician and primary care health care providers  Creating practice teams to implement the PEN model | PEN guidelines focused on lifestyle modification |  | Coordination between non-physician and physicians involved in delivering the PEN (Coordinated care) |
| Hirsch et al. (2017) |  | Organisation support the pharmacist-led MTM | Provision of patient-specific diabetes education | Support for pharmacist and endocrinologist activities related to education, collaborative care plan, and interpreting laboratory tests | MTM  Development of a personalised care plan using an MTM Spider Web model approach, which considers each patient’s comorbidities, complications, and clinical, socioeconomic, and behavioural issues |  | Coordination between pharmacist, endocrinologist, and other PCPs (coordinated care)  Personalised care plan developed using MTM spider web model approach that considers each patient’s comorbidities, complications, and clinical, socioeconomic, and behavioural issues (patient-centred care)  Provision of specific diabetes education during approximately three 60-minute visits over a 6-month period (Continuity of care) |
| Hobbs et al. (2005) |  | Organisation supports systematic screening of AF |  | GPs and practice nurses in the intervention practices received education on the importance of AF detection and ECG interpretation | Screening and education guidelines for the opportunistic screening program |  | Coordination between GPs and PNs for systematic screening of AF (Coordinated care) |
| Howard et al. (2010) |  | Organisations support Improved management of known patients with CKD risk factor and Primary care–based screening strategies for CKD risk factors |  | Support for providers to provide improved management and primary care-based screening for CKD risk factors |  |  | Intensive glucose and BP control (Patient-centred care) |
| Kasaie et al. (2020) | Multi Disease health campaigns under large tents in all communities during weekdays, evenings, and weekends in collaboration with local health units and the Ministry of Health in Uganda and in Kenya  Home-based testing for those who did not attend community campaigns | Program supports the community-based, multi-disease testing and treatment strategy |  | Support to healthcare providers to conduct multi disease health campaigns under large tents in all communities during weekdays, evenings, and weekends in collaboration with local health units and provide referral for patients found with HIV, hypertension, or diabetes | Use of the 2015 World Health Organization ART guidelines in which universal ART was recommended | In addition to residents’ names, biometric identifiers based on each resident’s digital fingerprint were used to identify residents during their participation in testing and care activities in the community. | Coordination between health facilities and mobile clinics (coordinated care)  Linkage to care for patients found to have HIV, hypertension, or diabetes (Continuity of care) |
| Kim et al. (2021) |  | Primary care clinics support the CDMP | Provision of health support services, such as professional health consultation and education | Support and incentives provided for health care providers participating in the CDMP |  |  | Patients receive continued care (continuity of care)  Provision of health support services, such as professional health consultation and education (Patient-centred care) |
| Mason et al. (2005) |  | Clinics supports the specialist nurse-led intervention to treat and control hypertension and hyperlipidemia in patients with diabetes | At subsequent visits, lifestyle factors were reinforced and reviewed, and medications were titrated according to response to the treatment and according to protocol. | Nurses received additional training in the management of hypertension and dyslipidemia in patients with diabetes from the local clinicians (J.M.G. and J.P.N.) and pharmacists | The use of existing guidelines to treat and control hypertension and hyperlipidemia in patients with diabetes |  | Coordination between nurses clinicians, and pharmacists (coordinated care)  Individualised action based on the hospital visit (Person-centred care) |
| Mousa et al. (2021) |  | Organisations support pharmacist-led care | Pharmacists provided medication counselling, offered instructions on self-monitoring, and advised patients on healthy lifestyle choices | Support for pharmacist-led care for patients with type 3 diabetes | Guidelines for medication prescription and procedures in patients with T2D |  | Coordination between pharmacists and other health care providers (Coordinated care)  Monitoring drug adherence (Continuity of care) |
| Penaloza-Ramos et al. (2016) |  | Health facility support the self-management program | Patients randomly assigned to self-management were trained to  self-monitor BP and to self-titrate their antihypertensive medication | Support to enable patient to communicate with family physician from home |  |  | Patients attended two or three sessions, each lasting around an hour and monitored monthly (Continuity of care) |
| Sando et al. (2020) |  | HIV clinics support the NCD screening program |  | Support to healthcare providers in HIV clinics to conduct NCD screening | The use of NCD screening guidelines |  | Management of patients with NCDs at the HIV clinic (Continuity of care) |
| Schaufler et al. (2010) |  | Facilities support the screening of T2D | Prevention of  T2D in subjects diagnosed with pre-diabetes  by intensive lifestyle intervention | Support from health facility to healthcare providers to conduct preventive screening for diabetes | The use of diabetes screening guidelines |  | Prevention of  T2DM in subjects diagnosed with pre-diabetes  either by intensive lifestyle intervention or with metformin (Continuity of care) |
| Schouten et al. (2010) |  | Clinics supported the quality-improvement collaborative | Health care providers trained on self-management support to patients | Health care providers were directed and supported to change professional performance and care organisation and introduce patient self-management and a system to register clinical parameters. | Materials and information (change package) about the structure of diabetes care, targets for glycaemic and cardiovascular risk control and therapy in a step-up regimen | System to register clinical parameters | The collaborative brings together and supports  multiprofessional diabetes teams from primary care and outpatient clinics (coordinated care) |
| Schuetz et al. (2013)* |  | Health care facilities/ organisations in the countries support the vascular disease health check interventions |  | The organisations and health care providers are supported appropriately to deliver the health checks to the population | Guidelines and standard operating procedures for health care providers exist and are used for the health checks |  |  |
| Schultz et al. (2021) |  | MTM clinic supports pharmacist-led services | Pharmacists provide adherence assistance for the patients | Support for the pharmacist-led activities in the MTM clinic |  | All visits are documented in the patient’s [electronic medical record](https://www.sciencedirect.com/topics/medicine-and-dentistry/electronic-patient-record), and medication changes are made in collaboration with the patient’s providers. | Monthly face-to-face visits to any patient in need of global medication and disease state assistance (Continuity of care)  The clinic provides personalised and evidence-based medication regimen, adverse event monitoring, adherence assistance, and care coordination (person-centred care) |
| Wang et al. (2006) | Involvement of community social workers in the programme | Organisation supports the multiple disease screening programme |  | Support to health care providers to screen for breast, colorectal and liver cancers, cervical and oral neoplasia, diabetes, hypertension, osteoporosis, and hyperlipidaemia | Use of screening guidelines |  | Coordination between healthcare providers participating in the Keelung programme (Coordinated care)  Referral to care for patients identified through the screening (Continuity of care) |

Notes: MTM, medication therapy management; PEN, Package of essential NCD interventions; PN, practice nurse, GP, General practitioner; AF, atrial fibrillation; CKD, chronic kidney disease; BP, blood pressure; HIV, human immunodeficiency virus; ART, antiretroviral therapy; CDMP, chronic disease management program; T2D, type 2 diabetes; * The intervention was simulated and therefore assumptions made regarding the integrated care components
