## Supplemental appendix table S4 for "Application of decision-analytic models to inform integrated care interventions for cardiometabolic multimorbidity: A systematic review"

**Table S4: Results of the decision-analytic models in the economic evaluations**

|  |  | **Costs and resource use** | | | **Outcomes/ benefits** | | | **Inc. Costs** | **Inc. Outcome** | **CE threshold** | **ICER (Base Case)** | **Conclusion** |
| --- | --- | --- | --- | --- | --- | --- | --- | --- | --- | --- | --- | --- |
| **Author (year)** | **Interventions** | **Currency** | **Intervention** | **Comparator** | **Measure** | **Intervention** | **Comparator** |  |  |  |  |  |
| Afzali et al. (2012) | High-level PN involvement | AUD ($) | $56,779 | $65,517 | QALYs | 6.8 (6.3–7.3) | 6.5 (5.9–7.0) | -$8,738 (-$12,522 to -$4,954) | 0.3 (0.2–0.4) | Not reported | Not reported | Cost-effective |
| Dukpa et al. (2014) | PEN program | BTN | BTN 205 735 | BTN 210 023 | DALYs averted | - | - | - | 0.038 | 159 168- 477 504 BTN/DALY averted | -112 906 | Very cost-effective |
|  | Universal screening | BTN | BTN 203 897 | BTN 210 024 | DALYs averted | - | - | - | 0.016 | 160 168- 477 504 BTN/DALY averted | -112 907 | Very cost-effective |
| Hirsch et al. (2017) | DIMM clinic (2 years) | USD ($) | $899,371 | $962,565 | QALYs | 97 | 96 | - | - | Not reported | -63,194/QALY | Cost-effective |
|  | DIMM clinic (5 years) | USD ($) | $2,137,659 | $2,272,572 | QALYs | 222 | 218 | - | - | Not reported | -33,728/QALY | Cost-effective |
|  | DIMM clinic (10 years) | USD ($) | $3,879,964 | $4,114,363 | QALYs | 385 | 375 | - | - | Not reported | -23,440/QALY | Cost-effective |
| Hobbs et al. (2005) | Opportunistic screening | GBP (£) | Not reported | Not reported | Cases detected | 75 (59–94) | 47 (35–62) | £10,174 (£9593 to £10,755) | 28 | Not reported | 363/ case detected | Cost-effective |
|  | Systematic high risk | GBP (£) | Not reported | Not reported | Cases detected | 53 (40–69) | 48 (35–62) | £24,530 (£23,678 to £25,382) | 6 | Not reported | Dominated by opportunistic screening | Systematic high risk: Dominated by opportunistic screening |
|  | Systematic population | GBP (£) | Not reported | Not reported | Cases detected | 74 (58–93) | 49 (35–62) | £48,260 (£46,952 to £49,567) | 27 | Not reported | Dominated by opportunistic screening | Systematic population: Dominated by opportunistic screening |
| Howard et al. (2010) | Primary care screening plus intensive treatment of diabetes | AUD ($) | $17,832 ($3,027–70,025 | $16,487 ($1,875–68,20 | QALYs | 12.798 (4.321–17.720) | 12.701 (4.144–17.627 | $1,345 (-$6,600–9,902) | 0.097 (–0.408–0.696 | $A50,000 | $13,866/QALY | Cost-effective |
|  | Primary care screening plus intensive treatment of hypertension | AUD ($) | $14,061 ($1,178–61,009) | $14,004 ($1,402–63,661) | QALYs | 12.947 (4.768–18.037 | 12.831 (4.673–17.69 | $57 (-$8,058–7,75 | 0.116 (–1.396–1.745) | $A50,000 | $491/QALY | Cost-effective |
|  | Primary care screening plus intensive treatment of protenuria | AUD ($) | $16,974 ($1,867–65,23 | $16,821 ($1,641–64,826 | QALYs | 12.763 (4.871–17.806) | 12.731 (4.828–17.806) | $153 (-$7,708–7,5 | 0.032 (–0.790–0.9 | $A50,000 | $4,781/QALY | Cost-effective |
| Kasaie et al. (2020) | Community-based integrated HIV-NCD screening and treatment | USD ($) | 860.36 [830.59 to 890.66] | - | DALYs averted | - | - | 6.68 [6.61 to 6.74] billion | 7.76 [8.01 to 7.51] million | USD 2,010 | $860.30 per DALY averted. | Cost-effective |
| Kim et al. (2021) | CDMP | Korean Won (KRW) | CDMP: 32 774 249 | Usual care: 35 473 613 | QALYs | 16.6 | 16.1 | Incremental: 2 699 364 | Incremental: -0.46855 | USD 33 429 | −5 761 088 (Usual care dominated by CDMP) | Highly Cost-effective |
| Mason et al. (2005) | Nurse-led clinics (BP control) | USD ($) | $306,400 | Not reported | QALYs | 0.53/patient | Not reported | - | - | $50,000/QALY | −$1,400/QALY | Cost-effective |
|  | Nurse-led clinics (Lipid lowering) | USD ($) | $306,400 | Not reported | QALYs | 0.46/patient | Not reported | - | - | $50,000/QALY | $8,230/QALY | Cost-effective |
| Mousa et al. (2021) | Pharmacist-led care | JD | 6668.5 (6244.29,7687.03) | 5391.2 (4931.6,6556.9) | Life years gained (LYG) | 4.2 (4.1,4.3) | 3.9 (3.8,4.0) | 1238.78 | 0.3 LYG/patient | (JD3,008.36) US$4,241.79 (very cost-effective) to (JD9,023.23) US$12,723 (cost-effective) | JD4058.5 (3208.8,6117.8) per LY gained | Cost-effective |
| Penaloza-Ramos et al. (2016) | Self-management | GBP (£) | 7357 | 8187 | QALYs | 6.2466 | 6.0326 | –830 | 0.2139 | £20,000/QALY gained | Self-management is Dominant | Cost-effective |
| Sando et al. (2020) | Integrated screeening and treatment (Women 60-69 years) | USD ($) | $10,541,000 | - | DALYs averted | 7305  - | - | - | - | $3,474 | 1445/ DALY averted | Cost-effective among older age-groups  Not cost-effective among 30-44 years age group |
|  | Integrated screeening and treatment (Men 60-69 years) | USD ($) | $2386000 | - | DALYs averted | 1700 | - | - | - | $3,474 | 1400/DALY averted | Cost-effective among older age-groups  Not cost-effective among 30-44 years age group |
| Schaufler et al. (2010) | Lifestyle intervention | Euro (€) | 24700 | 23000 | QALYs | - | - | €1637 | 2.91 | Not reported | €562.54 per QALY | Cost-effective in the longterm |
|  | Prevention with metformin | Euro (€) | 24000 | 23000 | QALYs | - | - | € 921 | 2.83 | Not reported | €325.44 per QALY | Cost-effective in the longterm |
| Schouten et al. (2010) | Quality improvement collaborative (Men) | Euro (€) | - | - | QALYs | - | - | € 860 | 0.33 | €20,000 to €80,000 per QALY gained | 2570/discounted QALY | Cost-effective |
|  | Quality improvement collaborative (Women) | Euro (€) | - | - | QALYs | - | - | € 643 | 0.26 | €20,000 to €80,000 per QALY gained | €2448/ discounted QALY | Cost-effective |
| Schuetz et al. (2013) | Standardized vascular disease health check | Euro (€) | Country specific costs not reported | Country specific costs not reported | QALYs | Country specific QALYs not reported. | Country specific QALYs not reported. | - | - |  | Cost per QALY gained (ICER) Denmark: 11595 France: 14903 Germany: 115 Italy: 11113 Poland: Cost saving UK: 2426 | Cost-effective in all study countries |
| Schultz et al. (2021) | MTM program | USD ($) | $1 378 052 | 0 | QALYs | 1402 | 1384 | $1 378 052 | 18 | $100,000 | $38 798/QALY | Cost-effective |
| Wang et al. (2006) | Multiple disease screening (100%) | USD ($) | 154,139,608 | 151,290,562 | Life years gained (LYG) | 854,143.59 | 849,871.29 | 4,272.30 | - | Not reported | US$667 | Multiple screening may be more cost-effective than single disease screening |

Notes: Base case results from selected studies are presented.
