## Supplemental appendix Section S1 for "Application of decision-analytic models to inform integrated care interventions for cardiometabolic multimorbidity: A systematic review"

**Appendix section S1: Search strategies for electronic databases**

1. **Medline search Strategy**

1	((effectiveness or cost effective* or microsim* or simulation or cost utility or cost-utility or cost minimi#ation or cost-minimi#ation or Markov or agent-based or mathematical or cost benefit or cost-benefit) adj model*).tw.

2	exp Models, Mathematical/

3	(decision adj1 (tree$ or analy$ or model$)).tw

4	exp "Costs and Cost Analysis"/

5	exp Cost-Benefit Analysis/

6	(economic adj (evaluation* or impact)).tw.

7	or/1-6

8	exp "Delivery of Health Care, Integrated"/

9	exp Comprehensive Health Care/

10	exp "Continuity of Patient Care"/

11	exp Patient-Centered Care/

12	exp "Referral and Consultation"/

13	(referral adj consultation).tw

14	((transmural or seamless) adj care).tw.

15	exp Patient Care Team/

16	((integrat* or coordinat* or horizontal or vertical) adj2 (care or service* or program* or deliver* or management)).tw.

17	(multi team or multiteam or multi care or multicare or multi clinic or multiclinic or multi service or multiservice or multi program* or multiprogram*or multi delivery or multidelivery or multi management).tw.

18	or/8-17

19	exp Chronic Disease/

20	(chronic adj2 (condition* or illness* or disease* or disorder*)).tw.

21	exp Noncommunicable Diseases/

22	non-communicable disease.mp

23	or/19-22

24	exp Metabolic Syndrome/ or exp Hypertension/ or exp Cardiovascular Diseases/ or exp Obesity/ or exp Metabolic Diseases/ or exp Diabetes Mellitus, Type 2/

25	kidney disease/

26	(Cardiovascular or cardio-vascular or CVD or heart disease* or cardiometabolic or cardio metabolic or stroke or cerebrovasc* or circulatory disease or myocardial infarction or arteriosclero* or atherosclero* or CIMT or (carotid adj2 (intima-media or intima media or IMT or plaque)) or blood pressure or hypertens* or pulse wave velocity or augmentation index or arterial stiffness or arterial stiffening or metabolic syndrome or diabetes or fasting glucose or insulin or cholesterol or lipid profile or lipid* or triglyceride* or obesity).tw.

27	or/24-26

28	and/7,18,23,27 not regression.tw. not qualitative.tw. not survey.tw.

29	limit 28 to english language (940 articles)

2. **Embase search Strategy 1974 to 2023 Week 49**

1	((effectiveness or cost-effective* or microsim* or simulation or cost utility or cost-utility or cost minimi#ation or cost-minimi#ation or Markov or agent-based or mathematical or cost benefit or cost-benefit) adj model*).tw.	115243

2	exp Mathematical model/	915951

3	exp "Costs and Cost Analysis"/	410272

4	exp Cost-Benefit Analysis/	95511

5	(economic adj (evaluation* or impact)).tw.	39528

6	(decision adj1 (tree$ or analy$ or model$)).tw.	38406

7	1 or 2 or 3 or 4 or 5 or 6	1478842

8	exp "Delivery of Health Care, Integrated"/	13747

9	exp Comprehensive Health Care/	6545478

10	exp "Continuity of Patient Care"/	1039565

11	exp Patient-Centered Care/	1039565

12	exp "Referral and Consultation"/	160588

13	(referral adj consultation).tw.	88

14	((transmural or seamless) adj care).tw.	462

15	exp Patient Care Team/	7175

16	((integrat* or coordinat* or horizontal or vertical) adj (care or service* or program* or deliver* or management)).tw.	24611

17	(multi team or multiteam or multi care or multicare or multi clinic or multiclinic or multi service or multiservice or multi program* or multiprogram*or multi delivery or multidelivery or multi management).tw.	882

18	8 or 9 or 10 or 11 or 12 or 13 or 14 or 15 or 16 or 17	6553871

19	exp Chronic Disease/	245087

20	(chronic adj2 (condition* or illness* or disease* or disorder*)).tw.	502381

21	exp Noncommunicable Diseases/	12057

22	non-communicable disease.mp. [mp=title, abstract, heading word, drug trade name, original title, device manufacturer, drug manufacturer, device trade name, keyword heading word, floating subheading word, candidate term word]	14157

23	19 or 20 or 21 or 22	670845

24	exp Metabolic Syndrome/ or exp Hypertension/ or exp Cardiovascular Diseases/ or exp Obesity/ or exp Metabolic Diseases/ or exp Diabetes Mellitus, Type 2/	7503141

25	kidney disease/	131142

26	(Cardiovascular or cardio-vascular or CVD or heart disease* or cardiometabolic or cardio metabolic or stroke or cerebrovasc* or circulatory disease or myocardial infarction or arteriosclero* or atherosclero* or CIMT or (carotid adj2 (intima-media or intima media or IMT or plaque)) or blood pressure or hypertens* or pulse wave velocity or augmentation index or arterial stiffness or arterial stiffening or metabolic syndrome or diabetes or fasting glucose or insulin or cholesterol or lipid profile or lipid* or triglyceride* or obesity).tw.	4363534

27	24 or 25 or 26	8792333

28	7 and 18 and 23 and 27	11882

29	limit 28 to (english language and embase and article)	4546

30	(7 and 18 and 23 and 27) not regression.tw. not qualitative.tw. not survey.tw.	5790

31	limit 30 to (english language and embase and article)	2328

3. **Web of Science Search strategy**

1	effectiveness model* or cost effective* model* or microsim* model* or simulation model* or cost utility model* or cost-utility model* or cost minimi$ation model* or cost-minimi$ation model* or Markov model* or agent-based model* or mathematical model* or cost benefit model* or cost-benefit model*

2	Cost Analysis

3	Cost-Benefit Analysis

4	economic NEAR (evaluation* or impact)

5	(decision NEAR (tree$ or analy$ or model$))

6	#5 OR #4 OR #3 OR #2 OR #1

7	Delivery of Health Care

8	Comprehensive Health Care

9	Continuity of Patient Care

10	Patient-Centered Care

11	referral NEAR consultation

12	(transmural or seamless) NEAR care

13	Patient Care Team

14	((integrat* or coordinat* or horizontal or vertical) NEAR (care or service* or program* or deliver* or management))

15	(multi team or multiteam or multi care or multicare or multi clinic or multiclinic or multi service or multiservice or multi program* or multiprogram*or multi delivery or multidelivery or multi management)

16	#7 OR #8 OR #9 OR #10 OR #11 OR #12 OR #13 OR #14 OR #15

17	Chronic Disease

18	(chronic NEAR (condition* or illness* or disease* or disorder*))

19	non-communicable disease

20	#17 or  #18 or #19

21	Metabolic Syndrome or Hypertension or Cardiovascular Diseases or Obesity or Metabolic Diseases or Diabetes*

22	kidney disease

23	(Cardiovascular or cardio-vascular or CVD or heart disease* or cardiometabolic or cardio metabolic or stroke or cerebrovasc* or circulatory disease or myocardial infarction or arteriosclero* or atherosclero* or CIMT or (carotid adj2 (intima-media or intima media or IMT or plaque)) or blood pressure or hypertens* or pulse wave velocity or augmentation index or arterial stiffness or arterial stiffening or metabolic syndrome or diabetes or fasting glucose or insulin or cholesterol or lipid profile or lipid* or triglyceride* or obesity)

24	#21 or #22 or #23

25	#6 AND #16 AND #20 AND #24 NOT regression or qualitative or survey

26	limit 25 to english language and article and Web of science core collection (2471 articles)

4. **Cochrane Library search strategy**

1	((effectiveness or cost effective* or microsim* or simulation or cost utility or cost-utility or cost minimi#ation or cost-minimi#ation or Markov or agent-based or mathematical or cost benefit or cost-benefit) NEAR model*)

2	(decision NEAR (tree# or analy* or model*))

3	Costs and Cost Analysis

4	Cost-Benefit Analysis

5	economic evaluation

6           economic impact

7	#1 OR #2 OR #3 OR #4 OR #5 OR #6

8	“Delivery of Health Care”

9	“Comprehensive Health Care”

10	“Continuity of Patient Care”

11	“Patient Centered Care”

12	"Referral and Consultation"

13	(referral adj consultation)

14	“transmural care”

15          “seamless care”

16	“Patient Care Team”

17	(integrat* or coordinat* or horizontal or vertical) NEAR (care or service* or program* or deliver* or management)

18	"multi team" or "multiteam" or "multi care" or "multicare" or "multi clinic" or "multiclinic " or "multi service" or "multiservice" or multi program* or multiprogram* or "multi delivery" or "multidelivery" or "multi management"

19	#8 OR #9 OR #10 OR #11 OR #12 OR #13 OR #14 OR #15 OR #16 OR #17 OR #18

20	Chronic Disease

21	(chronic NEAR (condition* or illness* or disease* or disorder*))

22	non-communicable disease

23	#20 or  #21 or #22

24	Metabolic Syndrome or Hypertension or Cardiovascular Diseases or Obesity or Metabolic Diseases or Diabetes*

25	kidney disease

26	(Cardiovascular or cardio-vascular or CVD or heart disease* or cardiometabolic or cardio metabolic or stroke or cerebrovasc* or circulatory disease or myocardial infarction or arteriosclero* or atherosclero* or CIMT or (carotid adj2 (intima-media or intima media or IMT or plaque)) or blood pressure or hypertens* or pulse wave velocity or augmentation index or arterial stiffness or arterial stiffening or metabolic syndrome or diabetes or fasting glucose or insulin or cholesterol or lipid profile or lipid* or triglyceride* or obesity)

27	#24 or #25 or #26

28	#7 AND #19 AND #23 AND #27 NOT regression or qualitative or survey (350 articles)

5. **CINAHL search strategy**

1	(effectiveness or cost effective* or microsim* or simulation or cost utility or cost-utility or cost minimi#ation or cost-minimi#ation or Markov or agent-based or mathematical or cost benefit or cost-benefit)

2	Mathematical model

3	Costs and Cost Analysis

4	Cost Benefit Analysis

5	economic evaluation

6           economic impact

7           (decision NEAR (tree# or analy* or model*))

8	#1 OR #2 OR #3 OR #4 OR #5 OR #6 OR #7

9	“Delivery of Health Care”

10	“Comprehensive Health Care”

11	“Continuity of Patient Care”

12	“Patient Centered Care”

13	"Referral and Consultation"

14	referral N2 consultation

15	“transmural care”

16          “seamless care”

17	“Patient Care Team”

18	(integrat* or coordinat* or horizontal or vertical) N2 (care or service* or program* or deliver* or management)

19	"multi team" or "multiteam" or "multi care" or "multicare" or "multi clinic" or "multiclinic " or "multi service" or "multiservice" or multi program* or multiprogram* or "multi delivery" or "multidelivery" or "multi management"

20	#9 OR #10 OR #11 OR #12 OR #13 OR #14 OR #15 OR #16 OR #17 OR #18 OR #19

21	Chronic Disease

22	(chronic NEAR (condition* or illness* or disease* or disorder*))

23	non-communicable disease

24	#21 OR #22 OR #23

25	Metabolic Syndrome or Hypertension or Cardiovascular Diseases or Obesity or  Metabolic Diseases or Diabetes*

26	kidney disease

27	(Cardiovascular or cardio-vascular or CVD or heart disease* or cardiometabolic or cardio metabolic or stroke or cerebrovasc* or circulatory disease or myocardial infarction or arteriosclero* or atherosclero* or CIMT or (carotid adj2 (intima-media or intima media or IMT or plaque)) or blood pressure or hypertens* or pulse wave velocity or augmentation index or arterial stiffness or arterial stiffening or metabolic syndrome or diabetes or fasting glucose or insulin or cholesterol or lipid profile or lipid* or triglyceride* or obesity)

28	#25 OR #26 OR #27

29	#8 AND #20 AND #24 AND #28 NOT regression or qualitative or survey

30          Limit to English (254 articles)

6. **APA PsychInfo via Ovid search strategy**

1	((effectiveness or cost effective* or microsim* or simulation or cost utility or cost-utility or cost minimi#ation or cost-minimi#ation or Markov or agent-based or mathematical or cost benefit or cost-benefit) adj model*).tw.

2	exp Models, Mathematical/

3	(decision adj1 (tree$ or analy$ or model$)).tw

4	exp "Costs and Cost Analysis"/

5	exp Cost-Benefit Analysis/

6	(economic adj (evaluation* or impact)).tw.

7	or/1-6

8	exp "Delivery of Health Care, Integrated"/

9	exp Comprehensive Health Care/

10	exp "Continuity of Patient Care"/

11	exp Patient-Centered Care/

12	exp "Referral and Consultation"/

13	(referral adj consultation).tw

14	((transmural or seamless) adj care).tw.

15	exp Patient Care Team/

16	((integrat* or coordinat* or horizontal or vertical) adj2 (care or service* or program* or deliver* or management)).tw.

17	(multi team or multiteam or multi care or multicare or multi clinic or multiclinic or multi service or multiservice or multi program* or multiprogram*or multi delivery or multidelivery or multi management).tw.

18	or/8-17

19	exp Chronic Disease/

20	(chronic adj2 (condition* or illness* or disease* or disorder*)).tw.

21	exp Noncommunicable Diseases/

22	non-communicable disease.mp

23	or/19-22

24	exp Metabolic Syndrome/ or exp Hypertension/ or exp Cardiovascular Diseases/ or exp Obesity/ or exp Metabolic Diseases/ or exp Diabetes Mellitus, Type 2/

25	kidney disease/

26	(Cardiovascular or cardio-vascular or CVD or heart disease* or cardiometabolic or cardio metabolic or stroke or cerebrovasc* or circulatory disease or myocardial infarction or arteriosclero* or atherosclero* or CIMT or (carotid adj2 (intima-media or intima media or IMT or plaque)) or blood pressure or hypertens* or pulse wave velocity or augmentation index or arterial stiffness or arterial stiffening or metabolic syndrome or diabetes or fasting glucose or insulin or cholesterol or lipid profile or lipid* or triglyceride* or obesity).tw.

27	or/24-26

28	and/7,18,23,27 not regression.tw. not qualitative.tw. not survey.tw.

29	limit 28 to english language (28 articles)

7. **Scopus search strategy**

1	effectiveness or cost effective* or microsim* or simulation or cost utility or cost-utility or cost minimi?ation or cost-minimi?ation or Markov or agent-based or mathematical or cost benefit or cost-benefit

2	Mathematical W/2 model

3	Costs and Cost Analysis

4	Cost Benefit Analysis

5	economic evaluation

6            economic impact

7            (decision W/2 (tree# or analy* or model*))

8	#1 OR #2 OR #3 OR #4 OR #5 OR #6 OR #7

9	“Delivery of Health Care”

10	“Comprehensive Health Care”

11	“Continuity of Patient Care”

12	“Patient Centered Care”

13	"Referral and Consultation"

14	referral W/2 consultation

15	“transmural care”

16          “seamless care”

17	“Patient Care Team”

18	(integrat* or coordinat* or horizontal or vertical) Pre/2 (care or service* or program* or deliver* or management)

19	"multi team" or "multiteam" or "multi care" or "multicare" or "multi clinic" or "multiclinic " or "multi service" or "multiservice" or multi program* or multiprogram* or "multi delivery" or "multidelivery" or "multi management"

20	#9 OR #10 OR #11 OR #12 OR #13 OR #14 OR #15 OR #16 OR #17 OR #18 OR #19

21	Chronic Disease

22	(chronic Pre/3 (condition* or illness* or disease* or disorder*))

23	non-communicable disease

24	#21 OR #22 OR #23

25	Metabolic Syndrome or Hypertension or Cardiovascular Diseases or Obesity or Metabolic Diseases or Diabetes*

26	kidney disease

27	(Cardiovascular or cardio-vascular or CVD or heart disease* or cardiometabolic or cardio metabolic or stroke or cerebrovasc* or circulatory disease or myocardial infarction or arteriosclero* or atherosclero* or CIMT or blood pressure or hypertens* or pulse wave velocity or augmentation index or arterial stiffness or arterial stiffening or metabolic syndrome or diabetes or fasting glucose or insulin or cholesterol or lipid profile or lipid* or triglyceride* or obesity)

28           (carotid pre/2 intima-media) or (carotid pre/2 intima media) or (carotid pre/2 IMT) or (carotid pre/2 plaque)

29	#25 OR #26 OR #27 OR #28

30	#8 AND #20 AND #24 AND #29 NOT regression or qualitative or survey

31	Limit #30 to article (714 articles)
