## Supplemental appendix Section S2 for "Application of decision-analytic models to inform integrated care interventions for cardiometabolic multimorbidity: A systematic review"

**Appendix section S2: Detailed description of the study selection process**

The study selection process commenced with the import of searched outputs into the Endnote citation manager, where duplicates were meticulously handled. Initial elimination prioritized duplicates based on titles, followed by a more thorough examination involving author names and publication year to ensure comprehensive duplication identification.

Post-duplicate removal, the articles were exported as an XML file into Covidence software, chosen for its user-friendly interface. Covidence facilitated efficient screening by streamlining the eligibility assessment process, promoting collaboration among reviewers, and ensuring transparency in study selection.

The next phase involved independent screening by four reviewers (EW, JO, CA), each utilizing a predefined selection checklist. Titles and abstracts were rigorously assessed, with those meeting the eligibility criteria proceeding to full-text screening. Throughout this process, reviewers remained blinded to each other’s decisions, enhancing the objectivity of the study selection. Any conflicts identified during screening were resolved through thorough discussion with a third reviewer (either of PD, RA, DG and PO), ensuring consistency and reliability.

The study selection adhered to predefined inclusion and exclusion criteria outlined in the systematic review protocol, contributing to the overall robustness of the systematic review. The PRISMA flow diagram in the main document visually represents the number of records identified, screened, assessed for eligibility, and ultimately included in the systematic review.

This comprehensive procedure, characterized by transparency and thoroughness, ensures the reliability and reproducibility of the study selection process.
