## Supplemental appendix Section S3 for "Application of decision-analytic models to inform integrated care interventions for cardiometabolic multimorbidity: A systematic review"

**Appendix section S3: Summary of results from the economic evaluations**

Except for one, all the included studies found integrated care to be cost-effective at the chosen willingness-to-pay threshold. The resulting incremental cost effectiveness ratio (ICERs) in a study in Uganda were higher than the chosen WTP ($3,474), but the integration of NCD services into existing HIV care was associated with decreased 10-year CVD risk and lower ICERs among the older age groups (60 to 69 year-olds) ^61^. In five studies conducted in Bhutan, USA, Korea, and UK, the analysis found that the integrated care intervention dominated the usual care alternative ^62,63,73,78,82^. In all the included studies, the sensitivity analyses matched the base-case analysis where integrated care remained cost-effective at different modelling scenarios used. Results from one-way sensitivity analysis of the studies indicated that the parameters with the greatest impact on ICERs were, intervention/ treatment effectiveness inputs ^61,63,76^, intervention costs e.g. reimbursement of staff, treatment costs ^61,76^, health state utilities ^85^, screening/ treatment coverage ^76^, time horizon ^77^. Cost-effectiveness acceptability curves (CEACs) were used in five studies ^75,77,79,81,82^ and showed cost-effectiveness of integrated care at a wide range of WTP thresholds.
